# Cognitive Behavioural Therapy in Adults with Psychogenic Non-Epileptic Seizures: A Systematic Review and Meta-Analysis of Randomised Controlled Trials

**DOI:** 10.1101/2024.05.10.24307189

**Authors:** Pierludovico Moro, Simona Lattanzi, Christoph P Beier, Carlo Di Bonaventura, Emanuele Cerulli Irelli

**Author notes:** **Corresponding author:** Carlo Di Bonaventura, MD, PhD, Department of Human Neurosciences, “Sapienza” University, P.le A. Moro, 5, Rome, Italy, 00185.

## Abstract

**Background:** Randomised controlled clinical trials (RCTs) investigating cognitive-behavioural therapy (CBT) among adults with psychogenic non-epileptic seizures (PNES) has become increasingly available, prompting the opportunity to critically appraise the efficacy and safety of CBT in this population.

**Methods:** We conducted a systematic review and meta-analysis including RCTs comparing CBT in addition to standardised medical treatment (SMT) versus SMT alone for adults with PNES. The primary outcome was seizure freedom at the end of treatment. Secondary outcomes included measures of quality of life, anxiety and depression assessed via standardised clinical questionnaires.

**Results:** Three RCTs were included comprising 228 participants treated with CBT and 222 with SMT. The intervention was significantly associated with seizure freedom (Odds Ratio [OR] 1.98; 95% confidence interval [CI] 1.14, 3.46; p = 0.02; I2 = 0%), reductions in anxiety (standardised mean difference [SMD] −0.21; 95% CI −0.41, −0.003; p = 0.047; I2 = 0%) and improvements in quality of life (SMD 0.34; 95% CI 0.12, 0.57; p = 0.003; I2 = 0%) at the end of treatment. Conversely, no significant differences between groups were observed regarding depression symptoms (SMD −0.19; 95% CI −0.39, 0.02; p = 0.08; I^2^ = 0%). There was no statistically significant increase in the risk of suicidal ideation and self-harm with CBT (OR 2.11; 95% CI 0.81, 5.48; p = 0.13; I^2^ = 0%) nor were there differences in terms of discontinuation rates during follow-up (OR 0.92; 95% CI 0.49, 1.72; p = 0.79; I^2^ = 7%).

**Conclusions:** There is high-quality evidence supporting the efficacy and safety of CBT in treating PNES. Future research should investigate whether combining CBT with other therapeutic methods could potentially enhance treatment efficacy.

## 1. Introduction

Psychogenic non-epileptic seizures (PNES) represent involuntary paroxysmal experiential and behavioural events that resemble epileptic seizures but do not coincide with the electroencephalography (EEG) changes observed in patients with epilepsy [1]. The reported estimated annual incidence of confirmed PNES is reported to be up to 4.90 cases per 100,000 persons. However, this incidence increases to 6.17 cases per 100,000 persons per year when including cases that are not confirmed by video-EEG [2].

The treatment of PNES has long been considered a challenge in clinical practice, particularly in adult patients with longer disease duration, who tend to have a poorer prognosis [3–6]. Various forms of psychotherapy administered by psychologists or psychiatrists have been proposed as the cornerstone of PNES treatment. Among these, cognitive-behavioural therapy (CBT) has emerged as a potential intervention over the last two decades [7]. Cognitive and behavioural therapies may be applied individually or combined within a comprehensive CBT approach. In cognitive therapy, the therapist helps the patient identify and correct distorted, maladaptive beliefs, while behavioural therapy employs learning principles to reduce symptoms and improve overall functioning, addressing both real and imagined stimuli [8, 9].

CBT is a well-established, evidence-based treatment for various psychiatric disorders, including depression, generalised anxiety disorder, posttraumatic stress disorder, panic disorder, eating disorders, substance use disorder, and obsessive-compulsive disorder [10]. Over the past decade, both observational and small randomised studies have suggested that CBT may potentially enhance neurological, psychiatric, and social outcomes in patients with PNES [11–23]. A prior meta-analysis, encompassing both randomised and non-randomized trials, indicated a reduction in PNES frequency of ≥50% following a comprehensive intervention involving various psychological treatments [24]. More recently, a large randomised controlled trial (RCT) investigating the role of CBT plus standard medical treatment (SMT) versus SMT alone in patients with PNES has been published [25], significantly expanding the population of randomised patients undergoing this intervention.

Given the availability of recent, high-quality randomised data, we conducted a systematic review and meta-analysis based exclusively on RCT data, aiming to evaluate the efficacy and safety of CBT in patients with PNES.

## 2. Methods

### 2.1 Search strategy

This systematic review and meta-analysis was registered a priori in the International Prospective Register of Systematic Reviews database (PROSPERO registration number: CRD42024512943). This study was reported in accordance with the Preferred Reporting Items for Systematic Reviews and Meta-analyses (PRISMA) reporting guidelines [26].

We conducted a comprehensive literature search using the MEDLINE, Cochrane, and Scopus databases electronic databases on November 28, 2023. The search strategy included synonyms and related terms based on the patient population (PNES OR psychogenic OR dissociative seizures OR functional seizures OR pseudoseizures OR pseudo seizures OR non epileptic attack disorder) and the intervention (psychotherapeutic Treatment OR psychoeducation OR cognitive behavioural therapy OR cognitive behavioural therapy OR cbt OR psychotherapy). The Boolean operators AND and OR were used to combine search terms and refine the search results. There were no date or language restrictions. The search strategy was adapted for each database as necessary (Supplement 1).

### 2.2 Literature Search and Study eligibility

Two investigators (P.M. and E.C.I.) independently appraised the titles and abstracts of the initial search results and selected studies for full-text review and screening. We included any RCT that compared CBT against SMT in adult patients (i.e., ≥18 years) diagnosed with PNES. We excluded non-original articles and grey literature or studies that did not undergo peer-review.

### 2.3 Outcome measures and data extraction

The primary outcome was seizure freedom at the end of treatment. Secondary outcomes included measures of quality of life, anxiety and depression assessed through standardised clinical questionnaires. The included studies utilised different standardised clinical questionnaires to quantitatively assess the variation in the analysed continuous outcomes. Specifically, for assessing QOL, the EuroQol Five-Dimensional Questionnaire (EQ-5D-5L) and the Quality of Life in Epilepsy Inventory 31 (QOLIE-31) were employed. For evaluating anxiety levels, the Generalised Anxiety Disorder seven-item scale (GAD-7), the Hospital Anxiety and Depression Scale (HADS), and the Beck Anxiety Inventory were utilised in the pooled analysis. Additionally, for measuring depression, the Patient Health Questionnaire nine-item scale (PHQ-9), the Beck Depression Inventory-II, and the HADS were considered.

As safety measures, we evaluated the occurrence of adverse events, including self-harm or suicidal ideation, and seizure worsening. Additionally, discontinuation rates between treatments were compared. Data was extracted and validated manually by two independent reviewers (P.M. and E.C.I.). In case of missing data, corresponding authors were contacted by email for requiring additional information.

### 2.4 Quality assessment

The methodological quality of the included studies was evaluated using the Cochrane risk-of-bias tool for randomised trials (RoB 2) [27]. Risk-of-bias evaluation was performed independently by two authors (P.M. and E.C.I.), with disagreements resolved by consensus of a third author (S.L.). Publication bias was assessed with funnel-plot analysis for the primary outcome of seizure freedom at the end of treatment.

### 2.5 Statistical analysis

Treatment effects for binary endpoints were evaluated using pooled odds ratios (ORs) along with 95% confidence intervals (CIs). Continuous endpoints were assessed through pooled standardised mean differences (SMDs) and standard deviations (SDs). The Mantel-Haenszel method was used for the binary endpoints and the Inverse Variance method was employed for the continuous endpoints. To assess heterogeneity, the Cochran’s Q test, I^2^ statistics, and Tau-square were employed, utilising the restricted maximum-likelihood estimator. Heterogeneity levels were categorised as low (I^2D^0–25%), moderate (I^2^L26–50%), or high (I^2^ >50%). Because of the potential methodological and sample differences, the random effects model was applied for all outcomes. This decision was made independently of heterogeneity considerations [28].

All statistical analyses were performed using RevMen Web [29]. The unavailability of raw data beyond median and interquartile range (IQR) values prevented the analysis of monthly seizure reduction. Attempts to transform these metrics into mean and standard deviation, following the methodologies proposed by Luo et al. and Shi et al. [30, 31], resulted in a highly skewed data distribution, which was not resolved through log transformation, preventing us from further analysis on this potential outcome analysis [32].

### 2.6 Sensitivity analysis

Leave-one-out sensitivity analysis was performed for the primary outcome to ensure the results were not dependent on a single study. This involved iteratively removing one study at a time to ensure the robustness of the results and to assess whether they were overly influenced by any single study.

## 3. Results

### 3.1 Study selection and baseline characteristics

The systematic search identified 2900 potential articles. Following the removal of duplicate records and the application of exclusion criteria based on title/abstract review, six articles remained and underwent thorough assessment for inclusion and exclusion criteria (Figure 1). One study was excluded due to persistent duplication [25], another was a study protocol [33], and one did not involve the intervention of interest [34]. Ultimately, three RCTs were included [18,23,25], encompassing a total of 450 patients. Among them, 228 patients were assigned to CBT, and 222 patients were assigned to SMT. Baseline characteristics of the participants of included studies are presented in Table 1. One RCT presented moderate risk of bias, whereas two other RCTs showed low risk of bias (Supplementary Figure 1).

**Figure 1.**
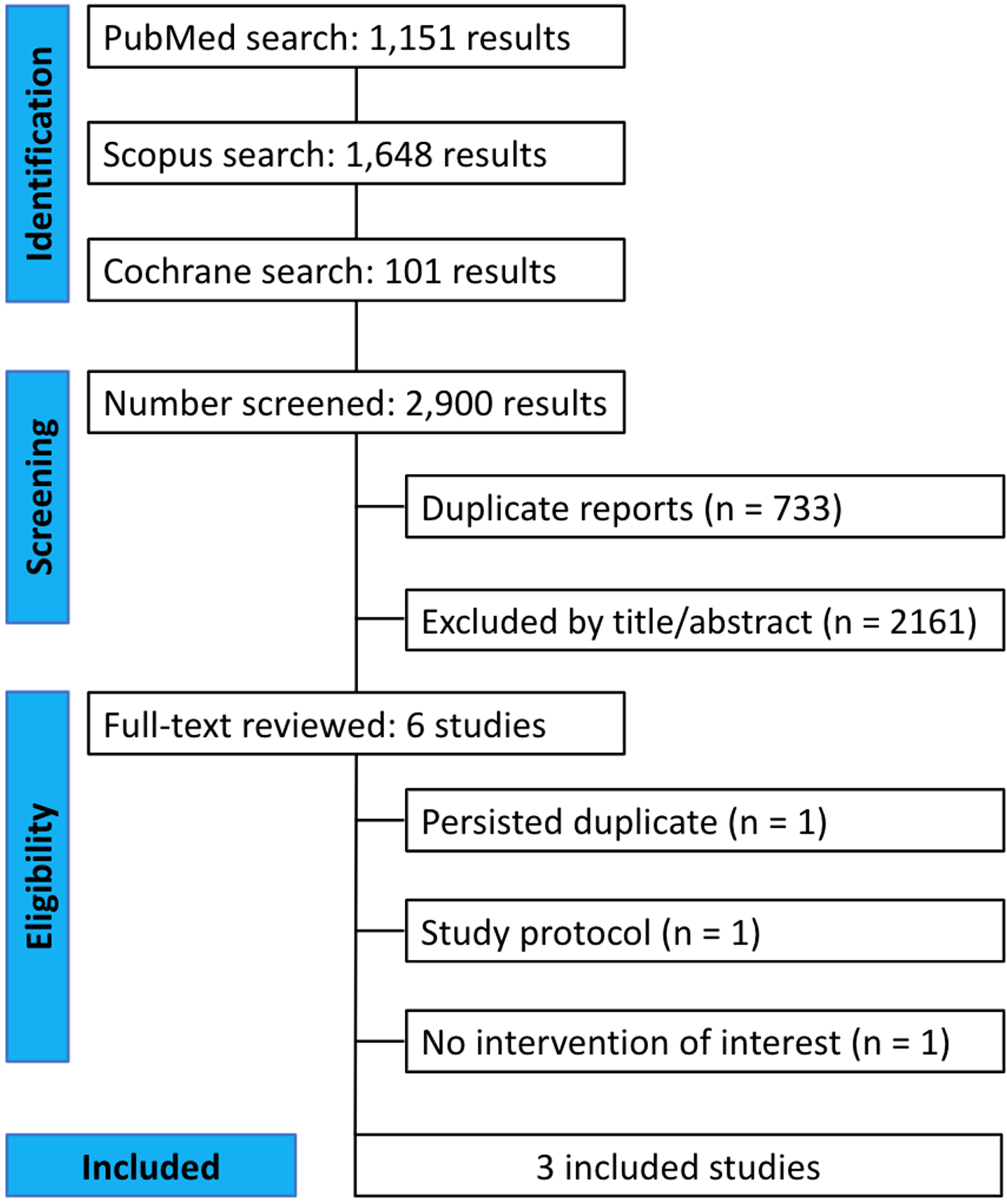
PRISMA flowchart presenting the selection of eligible studies.

**Table 1.**
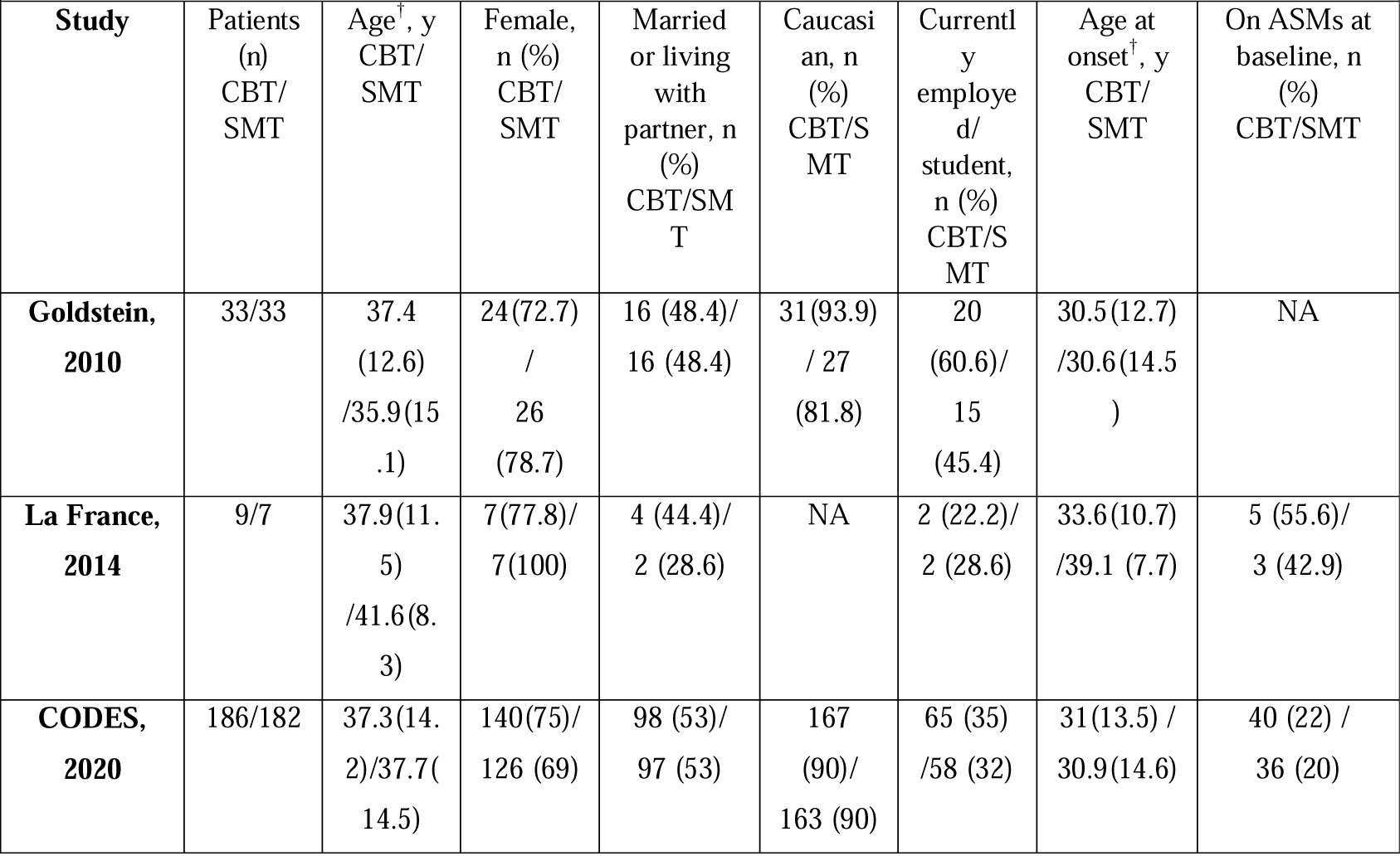
Design and characteristics of the studies included in the meta-analysis.

### 3.2 Primary outcome

Treatment with CBT was significantly associated with a higher likelihood of seizure freedom at the end of treatment (OR 1.98; 95% CI 1.14, 3.46; p = 0.02; I^2^ = 0%; Figure 2, panel A) compared to SMT. Given the variability in the duration of the intervention, despite the low heterogeneity of the primary outcome analysis, we conducted a leave-one-out sensitivity analysis, which provided similar results (Supplementary Figure 2, Panel A-C).

**Figure 2.**
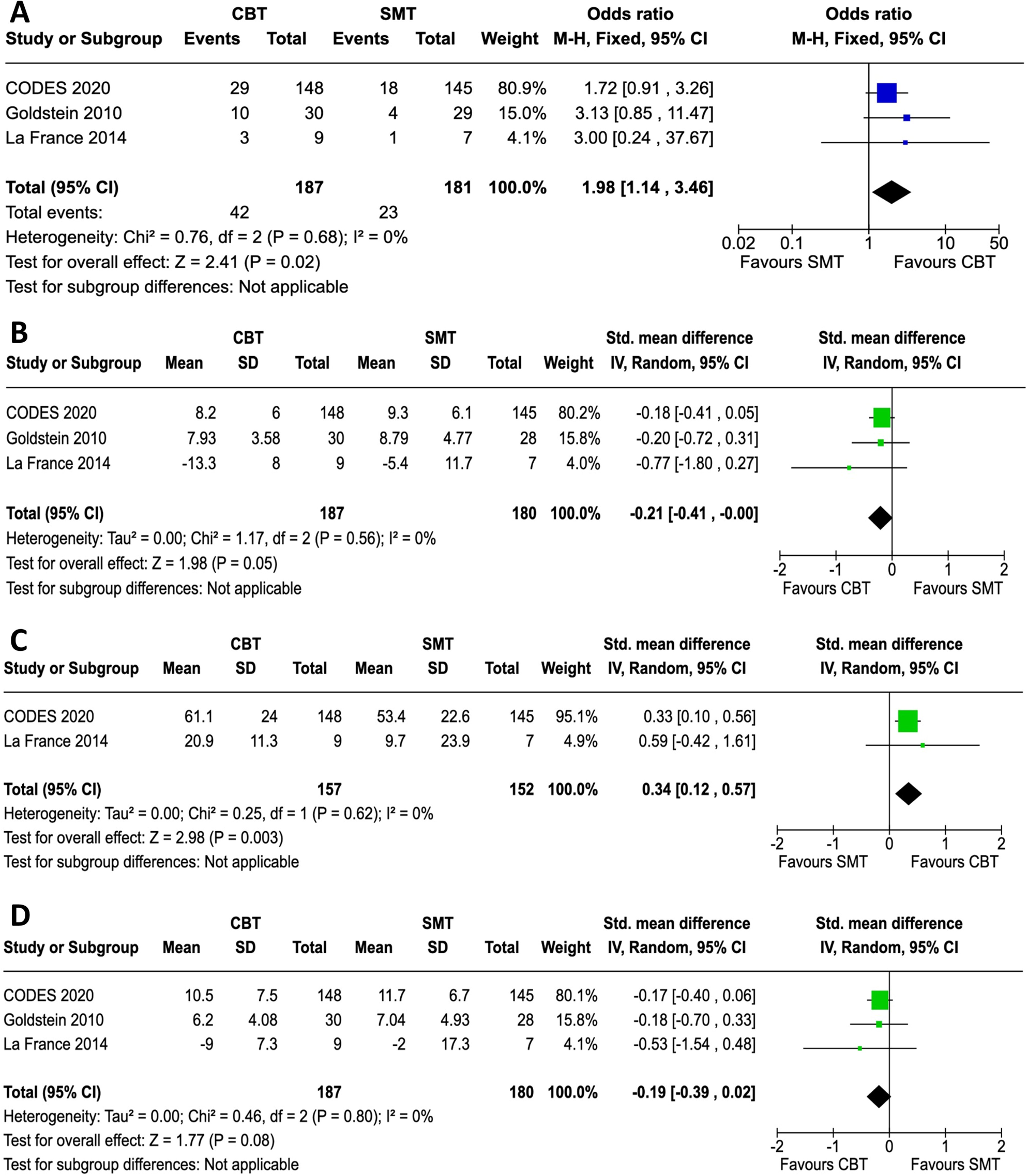
Analysis of efficacy outcomes. Forest plots presenting the association of CBT compared with SMT with seizure freedom (Panel A), anxiety (Panel B), quality of life (Panel C) and depression (Panel D) at the end of treatment. Abbreviations: CI, confidence interval; CBT, cognitive behavioural therapy; MH, Mantel-Haenszel; SD, standard deviation; SMT, standardised medical treatment; IV, inverse variance.

Visual examination of funnel plot revealed no apparent publication bias for the outcome of seizure freedom (Supplementary Figure 3).

### 3.3 Secondary outcomes

CBT significantly reduced anxiety (SMD −0.21; 95% CI −0.41, −0.003; p = 0.047; I^2^ = 0%; Figure 2, panel B) and improved quality of life (SMD 0.34; 95% CI 0.12, 0.57; p = 0.003; I^2^ = 0%; Figure 2, panel C) at the end of treatment compared with SMT alone. Conversely, no statistically significant differences between groups were observed in terms of depression symptoms (SMD −0.19; 95% CI −0.39, 0.02; p = 0.08; I^2^ = 0%; Figure 2, panel D) at the end of treatment.

### 3.4 Adverse events

Adverse events were reported in 2/3 RCTs (23, 25). Self-harm or suicidal ideation was reported in 6/215 (2.8%) participants receiving SMT and 13/219 (5.9%) patients treated with CBT, with no statistically significant difference between the two groups (OR 2.11; 95% CI 0.81, 5.48; p = 0.13; I^2^ = 0%) (Supplementary Figure 4, panel A). Worsening of seizure frequency was reported by only one study (18), in which an increase in seizure count during treatment was observed in 2/7 (28.6%) patients receiving SMT compared with 0/9 randomised to CBT. Finally, discontinuation rates during follow-up were not significantly different between CBT and SMT (OR 0.92, 95% CI 0.49-1.72, p=0.79, I^2^=7%) (Supplementary Figure 4, panel B). Reasons for discontinuation were not uniformly specified in included RCTs and were not considered in the meta-analysis.

## 4. Discussion

PNES present significant challenges in clinical management due to their multifaceted aetiology and clinical presentation. This systematic review and meta-analysis aimed to evaluate the efficacy and safety of CBT in adult patients with PNES, drawing on high-quality evidence from RCTs.

Our findings suggested that CBT may offer substantial benefits in the management of PNES. Specifically, treatment with CBT was associated with a higher likelihood of seizure freedom at the end of treatment compared to SMT alone. However, it is important to note that there was some degree of clinical heterogeneity in the measurement of seizure freedom among the included RCTs. This heterogeneity resulted from variations in the assessment of seizure freedom across trials; one RCT evaluated seizure freedom during the previous 3 months at a 12-month follow-up [25], another assessed it during the previous 3 months at a 6-month follow-up [23], while a third study observed seizure freedom over the entire 16-week observation period [18]. Despite this clinical heterogeneity, the leave-one-out sensitivity analysis provided similar results regarding the association of CBT with seizure freedom, validating the efficacy of this intervention.

In all three included RCTs, seizure reduction rather than seizure freedom was originally considered the primary outcome measure, and only two out of the three studies reported a significant reduction at the end of treatment [18, 23]. However, due to the highly skewed distribution of data, we were unable to pool the data for a meta-analysis. Nevertheless, it is worth noting that the use of mean seizure reduction as the main outcome measure has been questioned in both PNES and epilepsy literature due to its weak correlation with important measures of well-being and quality of life [35, 36], as well as difficulties in accurately calculating seizure frequency [37]. In contrast, seizure freedom represents a well-defined, more robust and crucial outcome in the management of both PNES and epilepsy [38], with several studies demonstrating its significant association with patient-reported outcome measures like quality of life to a greater extent than mere seizure reduction [39].

When considering secondary outcomes, this study revealed significant reductions in anxiety levels and improvements in quality of life among patients with PNES who received CBT. These results align with the broader literature demonstrating the effectiveness of CBT in addressing psychological distress and enhancing overall well-being in various clinical populations [8]. The observed reductions in anxiety highlight the potential of CBT to target and alleviate the increased levels of anxiety commonly experienced by individuals with PNES [40]. By helping patients to identify and challenge maladaptive thought patterns and unhelpful coping strategies, CBT provides them with the skills necessary to manage anxiety more effectively. Given that a large body of previous research has suggested that PNES may result from maladaptive coping strategies aimed at reducing anxiety and stress levels, the positive effect of CBT on anxiety levels might partially explain its observed efficacy in improving seizure freedom [41, 42].

Concurrently, the improvements in health-related quality of life among patients undergoing CBT highlight the broader impact of this therapeutic approach on the social and emotional aspects of PNES management. Quality of life encompasses various domains, including physical health, psychological well-being, social relationships, and overall life satisfaction [43]. The observed enhancements in quality of life suggest that CBT may not only alleviate seizure-related distress but also promote positive adjustments in other areas of patients’ lives.

However, it is noteworthy that this meta-analysis did not find a significant improvement in depressive symptoms among patients receiving CBT for PNES. While there was a small trend towards a positive effect on depressive symptoms, this finding did not reach statistical significance. When considering previous research, CBT represents a well-recognized and evidence-based treatment of major depression [44]. The observed discrepancy may reflect the complex interplay between depression and PNES, as well as the challenges inherent in treating comorbid psychiatric conditions within the context of PNES management. Additionally, the relatively small number of studies and heterogeneity among the measures used between studies may have limited the ability to detect significant effects on depressive symptoms.

Furthermore, this meta-analysis provided evidence regarding the safety of CBT in treating PNES, indicating no significant differences in adverse event rates. Additionally, CBT showed good tolerability and good adherence to treatment, with no differences in terms of discontinuation rates between CBT and SMT. However, it is noteworthy that adverse events were inconsistently reported in the included studies, despite the increasing documentation of adverse events during psychotherapeutic treatments in the literature [45]. Therefore, it would be pertinent for future RCTs to more accurately monitor the potential occurrence of adverse effects during CBT or other psychotherapeutic treatments. This monitoring should include self-harm or suicidal thoughts, which were slightly more frequent in the present study in patients receiving CBT compared to those receiving SMT, albeit at a non-statistically significant level.

This meta-analysis has some limitations. The included RCTs varied in methodology, treatment duration, and outcome measures, contributing to heterogeneity in the analyses. Additionally, the overall quality of evidence was moderate, with some concerns in the risk of bias. Finally, the inclusion of only three RCTs may limit the generalizability of the findings, as this number is considered the minimum for performing such analyses [28].

## 5. Conclusions

In conclusion, this systematic review with meta-analysis provided evidence supporting the efficacy of CBT as an adjunctive treatment for adult patients with PNES. Future research should focus on standardising CBT protocols, and explore whether multidisciplinary approaches, integrating CBT with other therapeutic methods, could provide more comprehensive care for individuals with PNES.

## Supporting information

Supplementary Figure 1. Risk of bias summary of randomized studies (RoB 2)

## Acknowledgements

None

## Ethics approval

Not applicable

## Patient consent for publication

Not applicable

## Data availability statement

All data relevant to the study are included in the article or uploaded as supplementary information.

## Abbreviations

CBT: Cognitive-Behavioural Therapy
CI: Confidence Interval
EEG: Electroencephalography
EQ-5D-5L: EuroQol Five-Dimensional Questionnaire
GAD-7: Generalised Anxiety Disorder seven-item scale
HADS: Hospital Anxiety and Depression Scale
OR: Odds Ratio
PHQ-9: Patient Health Questionnaire nine-item scale
PNES: Psychogenic Non-Epileptic Seizures
PRISMA: Preferred Reporting Items for Systematic Reviews and Meta-analyses
PROSPERO: International Prospective Register of Systematic Reviews
QOLIE-31: Quality of Life in Epilepsy Inventory 31
RCTs: Randomised Controlled Trials
RoB 2: Cochrane risk-of-bias tool for randomised trials
SD: Standard Deviation
SMD: Standardised Mean Difference
SMT: Standardised Medical Treatment

