## Supplementary Figure 1. Risk of bias summary of randomized studies (RoB 2) for "Cognitive Behavioural Therapy in Adults with Psychogenic Non-Epileptic Seizures: A Systematic Review and Meta-Analysis of Randomised Controlled Trials"

| Study | Bias from randomization process | Bias due to deviations from intended interventions | Bias due to missing outcome data | Bias in measurement of the outcomes | Bias in selection of the reported result | Overall risk of bias |
|---|---|---|---|---|---|---|
| CODES, 2020 | Low | Low | Low | Low | Low | Low |
| Goldstein, 2010 | Low | Low | Low | Low | Some concerns | Some concerns |
| La France, 2014 | Low | Low | Low | Low | Low | Low |

**Supplementary Figure 2. Leave-one-out sensitivity analysis of the primary outcome**

Each panel represents a separate analysis in which one of the randomized controlled trials has been excluded to assess the robustness of the results.

**A**

| Study or Subgroup | CBT Events | CBT Total | SMT Events | SMT Total | Weight | Odds ratio M-H, Random, 95% CI |
|---|---|---|---|---|---|---|
| Goldstein 2010 | 10 | 30 | 4 | 29 | 79.1% | 3.13 [0.85 , 11.47] |
| La France 2014 | 3 | 9 | 1 | 7 | 20.9% | 3.00 [0.24 , 37.67] |
| **Total (95% CI)** | | **39** | | **36** | **100.0%** | **3.10 [0.97 , 9.85]** |
| Total events: | 13 | | 5 | | | |

Heterogeneity: Tau² = 0.00; Chi² = 0.00, df = 1 (P = 0.98); I² = 0%
Test for overall effect: Z = 1.92 (P = 0.06)
Test for subgroup differences: Not applicable

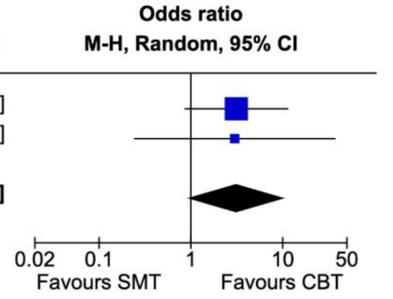


**B**

| Study or Subgroup | CBT Events | CBT Total | SMT Events | SMT Total | Weight | Odds ratio M-H, Random, 95% CI |
|---|---|---|---|---|---|---|
| CODES 2020 | 29 | 148 | 18 | 145 | 94.0% | 1.72 [0.91 , 3.26] |
| La France 2014 | 3 | 9 | 1 | 7 | 6.0% | 3.00 [0.24 , 37.67] |
| **Total (95% CI)** | | **157** | | **152** | **100.0%** | **1.78 [0.96 , 3.30]** |
| Total events: | 32 | | 19 | | | |

Heterogeneity: Tau² = 0.00; Chi² = 0.17, df = 1 (P = 0.68); I² = 0%
Test for overall effect: Z = 1.82 (P = 0.07)
Test for subgroup differences: Not applicable

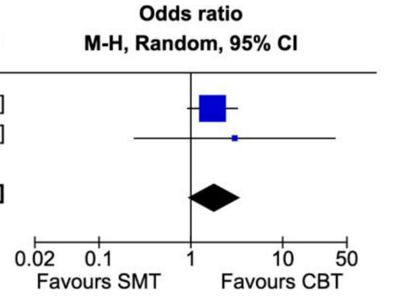


**C**

| Study or Subgroup | CBT Events | CBT Total | SMT Events | SMT Total | Weight | Odds ratio M-H, Random, 95% CI |
|---|---|---|---|---|---|---|
| CODES 2020 | 29 | 148 | 18 | 145 | 80.5% | 1.72 [0.91 , 3.26] |
| Goldstein 2010 | 10 | 30 | 4 | 29 | 19.5% | 3.13 [0.85 , 11.47] |
| **Total (95% CI)** | | **178** | | **174** | **100.0%** | **1.93 [1.09 , 3.43]** |
| Total events: | 39 | | 22 | | | |

Heterogeneity: Tau² = 0.00; Chi² = 0.65, df = 1 (P = 0.42); I² = 0%
Test for overall effect: Z = 2.25 (P = 0.02)
Test for subgroup differences: Not applicable

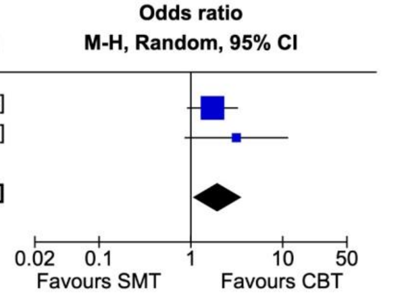

**Supplementary Figure 3. Funnel plot**

Publication bias for the binary outcome of seizure freedom at the end of treatment was evaluated through visualization of funnel-plot. The observed asymmetry, where the three included studies displayed varying weights on either side of the pooled effect line, was interpreted with caution due to the limited number of studies. Factors such as methodological differences or true heterogeneity might contribute to this pattern.

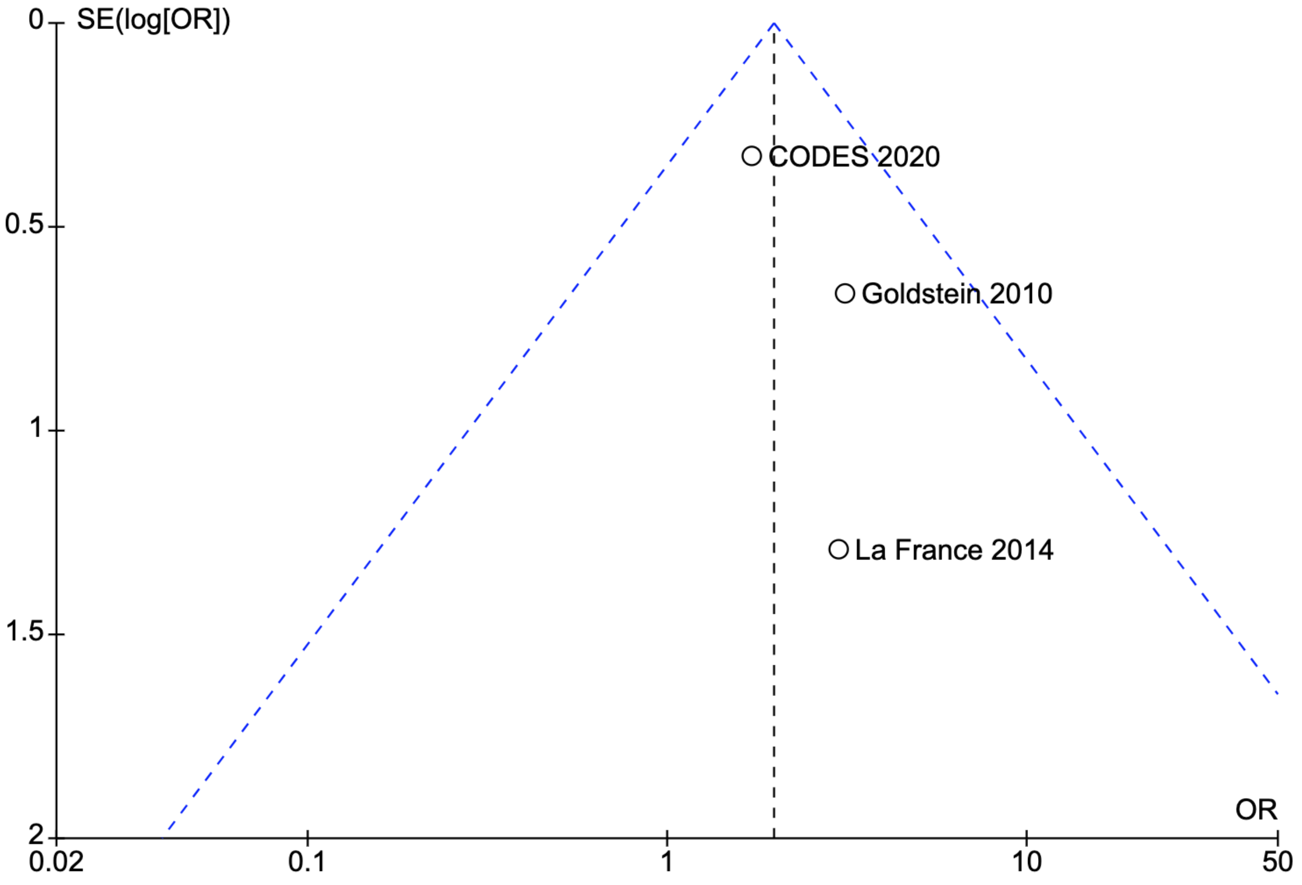

**Supplementary Figure 4. Forest plot of adverse events (self-harm or suicidal ideation, panel A) and discontinuation rates (Panel B)**

**A**

| Study or Subgroup | CBT Events | CBT Total | SMT Events | SMT Total | Weight | Odds ratio M-H, Fixed, 95% CI |
|---|---|---|---|---|---|---|
| CODES 2020 | 12 | 186 | 6 | 182 | 92.2% | 2.02 [0.74 , 5.51] |
| Goldstein 2010 | 1 | 33 | 0 | 33 | 7.8% | 3.09 [0.12 , 78.70] |
| **Total (95% CI)** | | **219** | | **215** | **100.0%** | **2.11 [0.81 , 5.48]** |
| Total events: | 13 | | 6 | | | |

Heterogeneity: Chi² = 0.06, df = 1 (P = 0.81); I² = 0%
Test for overall effect: Z = 1.53 (P = 0.13)
Test for subgroup differences: Not applicable

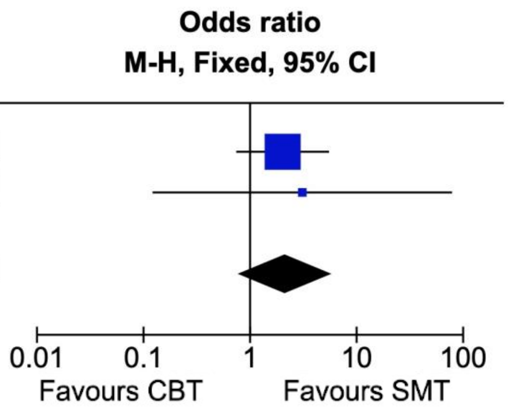


**B**

| Study or Subgroup | CBT Events | CBT Total | SMT Events | SMT Total | Weight | Odds ratio M-H, Random, 95% CI |
|---|---|---|---|---|---|---|
| CODES 2020 | 25 | 140 | 30 | 181 | 79.5% | 1.09 [0.61 , 1.96] |
| Goldstein 2010 | 3 | 30 | 4 | 29 | 14.7% | 0.69 [0.14 , 3.41] |
| La France 2014 | 1 | 9 | 3 | 7 | 5.9% | 0.17 [0.01 , 2.16] |
| **Total (95% CI)** | | **179** | | **217** | **100.0%** | **0.92 [0.49 , 1.72]** |
| Total events: | 29 | | 37 | | | |

Heterogeneity: Tau² = 0.04; Chi² = 2.15, df = 2 (P = 0.34); I² = 7%
Test for overall effect: Z = 0.27 (P = 0.79)
Test for subgroup differences: Not applicable

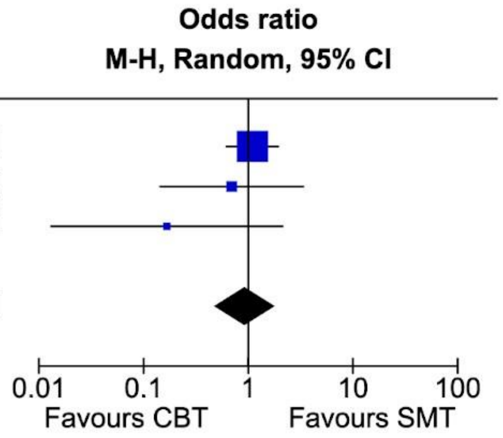
